# Integrated metabolic and genetic analysis reveals distinct features of primary differentiated thyroid cancer and its metastatic potential in humans

**DOI:** 10.1101/2023.03.09.23287037

**Authors:** Eduardo Cararo-Lopes, Akshada Sawant, Dirk Moore, Hua Ke, Fuqian Shi, Saurabh Laddha, Ying Chen, Anchal Sharma, Jake Naumann, Jessie Yanxiang Guo, Maria Gomez, Maria Ibrahim, Tracey L Smith, Gregory M. Riedlinger, Edmund C. Lattime, Stanley Trooskin, Shridar Ganesan, Xiaoyang Su, Renata Pasqualini, Wadih Arap, Subhajyoti De, Chang S. Chan, Eileen White

## Abstract

Differentiated thyroid cancer (DTC) affects thousands of lives worldwide every year. Typically, DTC is a treatable disease with a good prognosis. Yet, some patients are subjected to partial or total thyroidectomy and radioiodine therapy to prevent local disease recurrence and metastasis. Unfortunately, thyroidectomy and/or radioiodine therapy often worsen(s) the quality of life and might be unnecessary in indolent DTC cases. This clinical setting highlights the unmet need for a precise molecular diagnosis of DTC, which should dictate appropriate therapy. Here we propose a differential multi-omics model approach to distinguish normal gland from thyroid tumor and to indicate potential metastatic diseases in papillary thyroid cancer (PTC), a sub-class of DTC. Based on PTC patient samples, our data suggest that elevated nuclear and mitochondrial DNA mutational burden, intratumor heterogeneity, shortened telomere length, and altered metabolic profile reflect the potential for metastatic disease. Specifically, normal and tumor thyroid tissues from these patients had a distinct yet well-defined metabolic profile with high levels of anabolic metabolites and/or other metabolites associated with the energy maintenance of tumor cells. Altogether, this work indicates that a differential and integrated multi-omics approach might improve DTC management, perhaps preventing unnecessary thyroid gland removal and/or radioiodine therapy. Well-designed, prospective translational clinical trials will ultimately show the value of this targeted molecular approach.

**TRANSLATIONAL RELEVANCE:** In this article, we propose a new integrated metabolic, genomic, and cytopathologic methods to diagnose Differentiated Thyroid Cancer when the conventional methods failed. Moreover, we suggest metabolic and genomic markers to help predict high-risk Papillary Thyroid Cancer. Both might be important tools to avoid unnecessary surgery and/or radioiodine therapy that can worsen the quality of life of the patients more than living with an indolent Thyroid nodule.

**Graphical Abstract:** 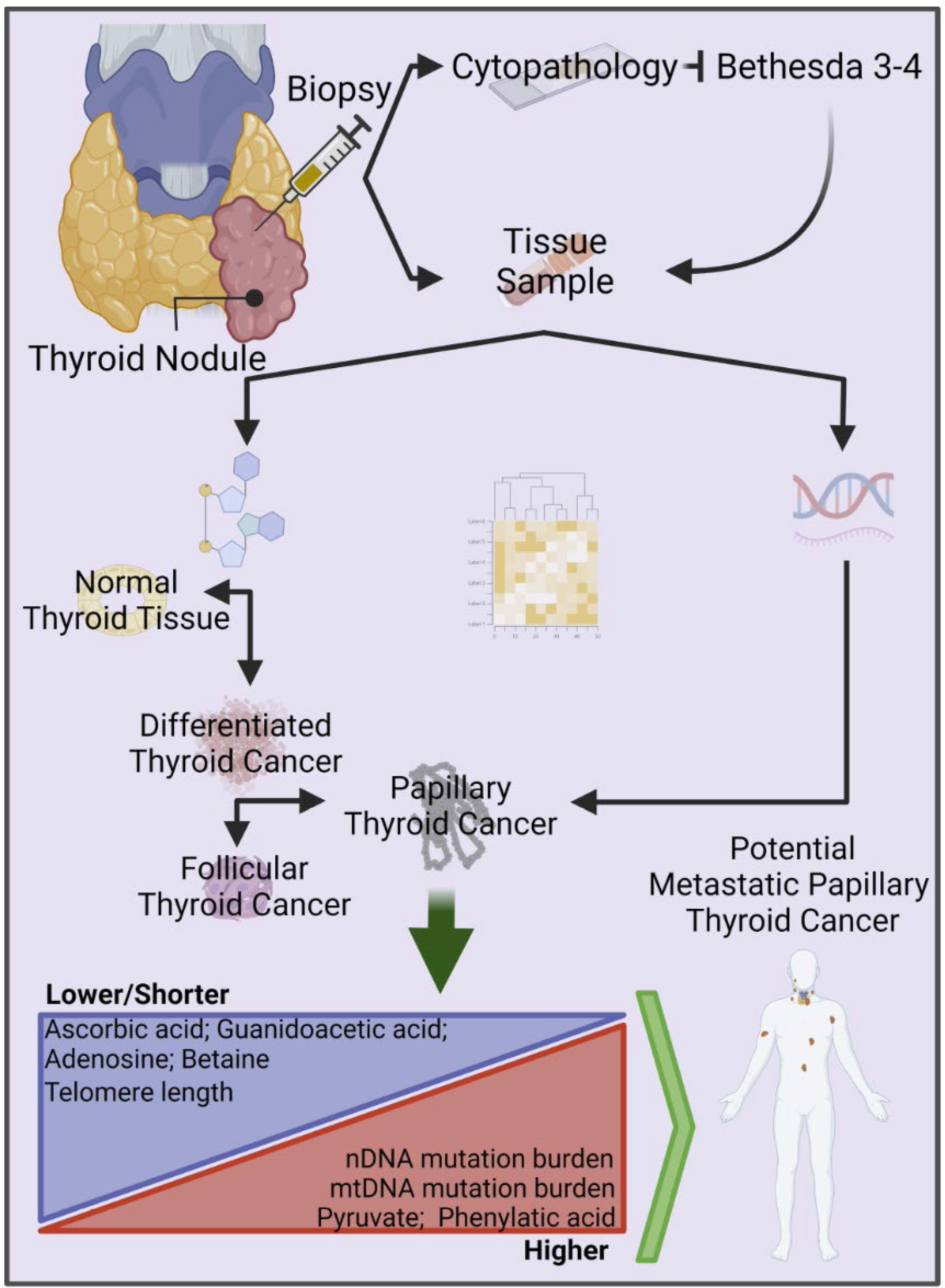

## INTRODUCTION

In 2021 the Surveillance, Epidemiology, and End Results (SEER) database estimated that thyroid cancer represents ∼2.3% of all new cancer cases in the United States and is responsible for ∼2.200 deaths in the same period. Follicular thyroid and parafollicular C cells are the two endocrine cells from which thyroid cancers putatively originate. However, thyroid tumors with the largest worldwide incidence arise from follicular cells. They are generically classified as differentiated thyroid cancer (DTC), which is divided into two histopathologic categories, namely: papillary thyroid cancer (PTC) and follicular thyroid cancer (FTC) (1, 2).

Currently, one concern related to DTC is its unexplained increase in the past 40 years. Although the incidence of DTC has been growing, the mortality of DTC patients has paradoxically remained stable (3, 4). Some investigators suggested that overdiagnosis (caused by the increased ability to detect indolent nodules and papillary cancers that would never cause symptoms or be life-threatening) may play a role (5, 6). However, the increased incidence of nodules larger than 5 cm is a genuine concern (7), as they are generally symptomatic and must be treated by surgical removal of the thyroid gland and, if cancer is present, both by adjuvant radioiodine therapy in certain cases (8, 9). Moreover, the epidemiologic data suggest that both the overdiagnosis and truly increased incidence of clinical thyroid nodules occur concomitantly, suggesting that a more accurate methodology is desirable in this setting.

Finally, an increasing number of poorly diagnosed thyroid cancer patients undergo surgical removal of part or the entire thyroid gland (8), which leads to several problems, such as inappropriate/unnecessary surgical or radioiodine therapy (9–11).

Therefore, the diagnostic method for differentiating benign from malignant tumors and indolent vs. aggressive thyroid cancers must be improved (12). The current gold standard procedure to diagnose thyroid cancer is still the cytopathologic analysis of thyroid nodule samples obtained from percutaneous fine-needle aspiration (FNA) by using the Bethesda Classification System (8, 13, 14). Unfortunately, FNA with pathologic analysis is inconclusive in up to ∼25%, bringing into question the choice of therapy (10, 15, 16). In order to overcome this challenge, molecular testing towards the precise diagnosis of indeterminate thyroid nodules were developed. The most used platforms are RNA-expression based detection (Veracyte;17) or identification of somatic mutations (ThyroSeq;18). Nevertheless, both methods also show their limitations (19) which might be complemented by new methods of diagnosis such as metabolic profiling.

On a further level of complexity, a small percentage of these tumors belong to high-risk thyroid cancer variants that often generate distant metastasis, with a very poor prognosis compared to non-metastatic thyroid cancer (20); also, up to 20% of patients with DTC may have residual tumors left behind after surgical or radioactive iodine treatment (1, 21–24). Therefore, there is a clear need for improved procedures and biomarkers to distinguish among benign nodules, low-risk and, high-risk thyroid cancer.

Here we evaluate the metabolomic profile of normal gland and thyroid tumors from DTC patients (n=20) with different features (e.g., gender, stage, metastatic status), showing that the metabolic profile of DTC tumor cells remains consistent and well-defined and may potentially be used to distinguish normal thyroid gland from cancer. Moreover, matched tissues from these patients (n=10), including normal and tumor samples, were subjected to whole-genome sequencing (WGS). Unexpectedly the WGS results indicate other features beyond the mutational profile, such as mitochondrial and nuclear DNA (mtDNA and nDNA) mutational burden and telomere length, may also serve as candidates biomarkers for metastatic PTC, including local lymph node metastasis (LM) and high-risk PTC presenting with distant metastases (DM).

On another note, we also compared the intra-tumor heterogeneity among multiple samples from the same thyroid tumor. We observed that histopathological heterogeneity strongly correlates to post-transcriptional events (such as gene expression and metabolic rewiring) independently of the conserved mutational profile. Together, these results highlight the translational importance of integrated omics for thyroid cancer diagnosis, prognosis, and management.

## RESULTS

### Differentiated thyroid cancer has a well-defined metabolic profile

In order to evaluate the metabolomic profile of differentiated thyroid cancer (DTC), we used normal tissue and primary thyroid tumor obtained from partial or total surgical thyroid resections (Fig. 1A). The histologic subtype was classified, and only the patients with DTC were selected. This cohort comprises 20 patients, 18 of these DTC patients (95%) have their tumors classified as papillary thyroid cancer, while the other two were follicular thyroid cancer (Supplementary Table 1). Most of the patients were women (75%) and, on average, were slightly younger than the men (Figs. 1B and 1C). Although the average size of the primary tumor in both genders was similar, in this series, the percentage of metastatic DTC samples (including distant and local metastasis) was higher in men, which may suggest a more aggressive disease, late clinical diagnosis, or both (Figs. 1D, 1E, and 1F). The cohort of patients evaluated here is generally representative of the natural history of DTC.

**Figure 1:**
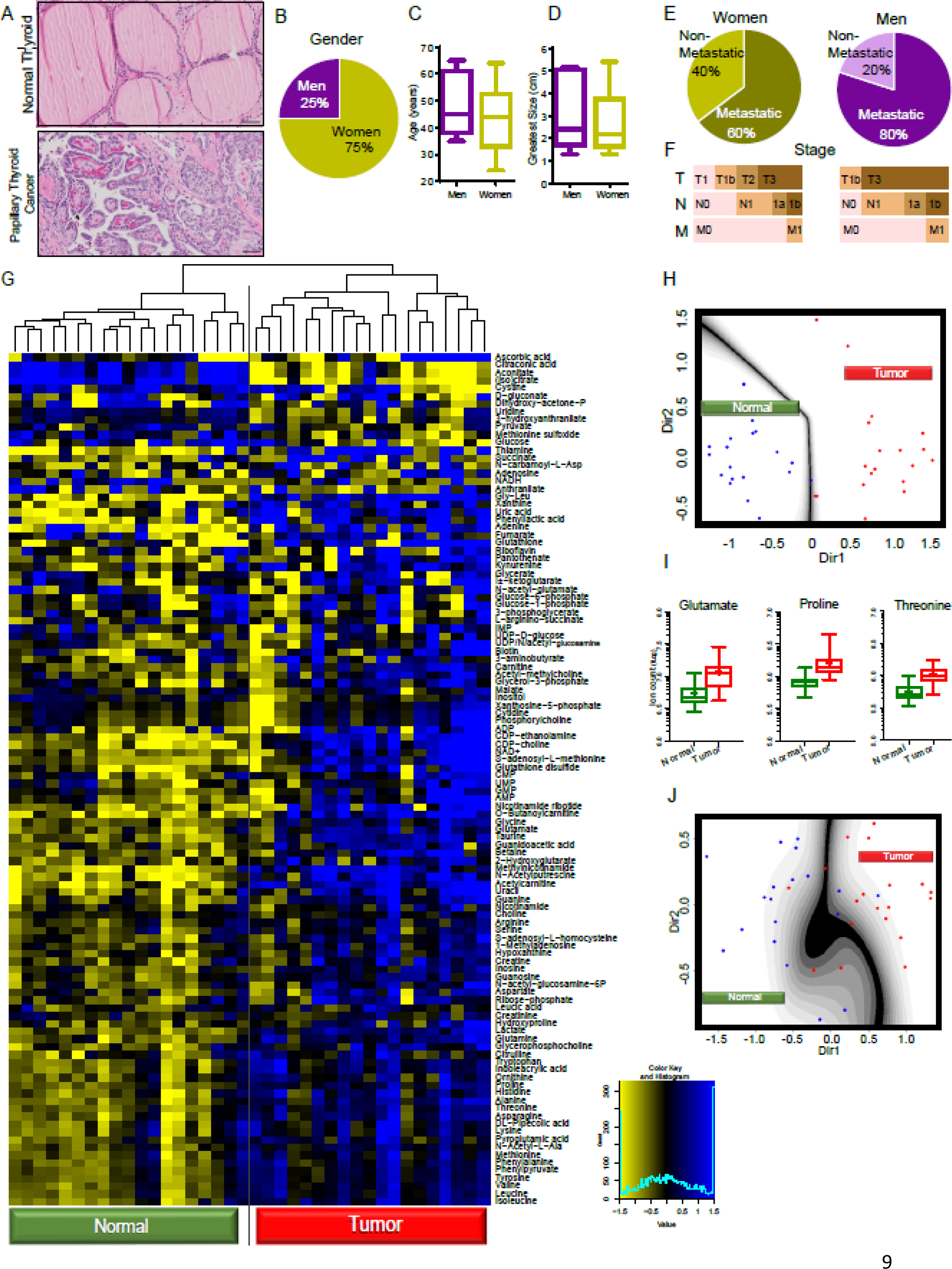
Differentiated thyroid cancer presents a well-defined metabolic profile. (A) H&E scanned slides of normal thyroid tissue and papillary thyroid cancer (Scale bar 50 µm). DTC patients’ profiles consider (B) gender, (C) age, and (D) the greatest size of the primary DTC tumor. (E) Thyroid cancer risk, metastatic (local and distant metastasis), and non-metastatic, per gender. (F) Classification of thyroid cancer according to histopathologic analysis in surgical report. Primary tumor (T), local metastasis (Lymph nodes (N)), and distant metastasis (other organs (M)). (G) Heat map of metabolites extracted from primary thyroid cancer and normal thyroid tissue. (H) mClust analysis considering the six most significant altered metabolites. All the points except one lie on their respective sides. That one, however, lies within the boundary error region. (I) Box plot of the three metabolites with lower p-value used to perform mClust analysis (J) with the lower number of biomarkers possible.

One of our goals was to identify metabolic biomarkers predictive of whether or not a tissue sample is normal or a tumor of the thyroid tissue. We also evaluated whether the metabolic changes might shed light on the molecular mechanisms of metastatic DTC. We focused on polar metabolites (n=110) from each sample. These analyses clearly resulted in a DTC metabolic profile distinct from normal thyroid (Fig. 1G). Next, we aimed to identify an informative subset of biomarkers, which may effectively differentiate the two types of tissue (normal vs. cancer) based on their metabolic profile. As a first step, we constructed a predictive model which uses ordinary logistic regression to assess the relationship of each compound to the sample type. We selected the metabolic biomarkers with *p*-values ≤ 0.20/110 = 0.0018, thereby practically reducing the number of candidate biomarkers (from 110 to 19). We next used cross-validated Lasso logistic regression by using the “optL1” function in the R package “penalized” to find the penalty parameter that minimizes the 10-fold cross-validated penalized likelihood (25). We then used the “penalized” R function in this package with this penalty parameter to identify the optimal subset of metabolic biomarkers (n=6) that were most effective in predicting normal versus cancer. The six biomarkers found were aconitate, glycine, inosine, isoleucine, proline, and taurine (Supplementary Fig. S1A). These results clearly show that applying the aforementioned predictive model in the metabolomic analysis distinguishes normal thyroid tissue from primary thyroid tumors.

To illustrate the ability to use these six compounds to separate normal and cancer samples, we next used a procedure known as “mclust” that identifies the projection of the six-dimensional predictors onto two dimensions that optimally separate the two groups (26, 27) (Fig. 1H). These results suggest that the six predictors selected may differentiate normal biopsy samples from thyroid tumors. To better evaluate the classification error, we applied the ten-fold cross-validation methods, which yielded an error rate of 0.1316 and a standard error of 0.0451, clearly differentiating this error rate from the null hypothesis value of 0.5 (Supplementary Fig. S1B).

Moreover, we considered whether an even smaller subset of biomarkers could be suitable for adequate classification. To this end, we performed the same procedure with five, four, or three of the most effective metabolic biomarkers in predicting the phenotype. However, the model with fewer predictors has lower power and many misclassified samples (Supplementary Figs. S1C and S1D), but in all cases, the error rates are below 0.5. For instance, selecting the three biomarkers (i.e., glutamate, proline, and threonine) (Fig. 1I) generated only three misclassifications per sample type, all lying in the error boundary (Fig. 1J). Applying ten-fold cross-validation, we could rely on an error rate of 0.21 and a standard error of 0.051, suggesting that this more straightforward procedure could be helpful in the absence of a more extensive metabolic panel. Notably, all proteogenic amino acids are present at higher levels in tumor samples (Supplementary Fig. S1E), making the classification procedure even more accessible. Thus, our data indicate that a histopathologic analysis supplemented by a small metabolomic biomarker panel might efficiently improve DTC diagnosis.

### The metabolic alterations of DTC are enriched in energy maintenance and anabolic metabolism

A volcano plot showed that most metabolites are present at higher levels in DTC (Fig. 2A). As 45 of 110 metabolites (41%) were significantly altered (adjusted *p-value* ≤ 0.01/110), we applied a more restrictive analysis considering only the metabolites that have fold change more than double, or less than half, in normal thyroid *versus* cancer tissue (Figs. 2A and 2B). Thus, we found the most affected metabolite classes and the pathways these biochemical alterations impact.

**Figure 2:**
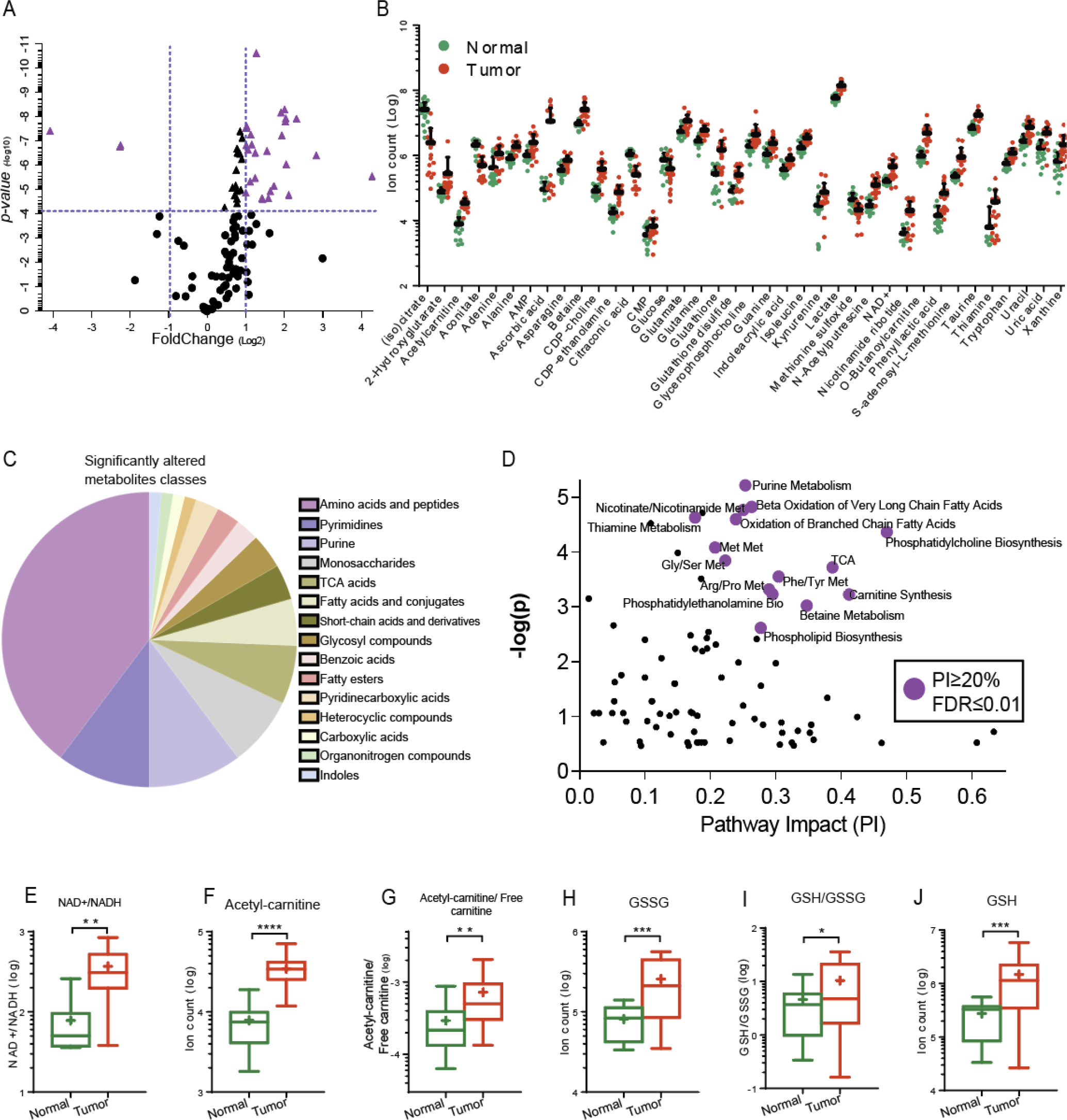
DTC prioritized metabolism of building blocks and energy maintenance. (A) Volcano plot of metabolites extracted from DTC patients’ samples. The purple triangles highlight the significantly altered metabolites (*p-value* ≤ 0.01/110) with fold change higher than double or less than a half comparing cancer versus normal thyroid tissues. The levels of these metabolites are graphically expressed in (B). Data presented as mean (SEM). (C) The most altered classes of metabolites in DTC. (D) Pathways impact analysis. Purple circles show the metabolic pathways impacted in more than 20%, with a false discovered ratio equal to or lower than 0.01. Box plot of metabolites involved in energy maintenance (E-G) and oxidative stress (H-J). T-test ** *P*≤0.01.

According to this stringent criterion, ∼70% of the metabolites are amino acids, purines, pyrimidines, 1-Carbon metabolism intermediates, tricarboxylic acid (TCA) cycle components, and fatty acids (Fig. 2C). The most altered metabolites in malignant DTC are involved in two fundamental processes: metabolism of building blocks and energy maintenance. Indeed, the pathway impact analysis revealed that the main altered pathways in DTC are the TCA cycle, beta-oxidation of fatty acids, amino acids, and purine/pyrimidine metabolism (Fig. 2D). A high-energy requirement of thyroid tumor cells might be responsible for the altered TCA intermediate metabolites, as suggested by the higher NAD+/NADH ratio in tumors (Fig. 2E). However, the most robust result linked to energy maintenance was the increased levels of several metabolites from lipid metabolism. For instance, a lower level of free carnitine in tumor cells may be one example because one of its main metabolic roles is to shuttle long-chain fatty acids across the mitochondrial membrane to be burned by β-oxidation (Fig. 2G). Furthermore, the higher acetyl-carnitine/free carnitine ratio also suggests that DTC might have altered β- oxidation activity, consistent with previous observations (28).

As with other malignant tumor cells, thyroid cancer cells are driven to increase the proliferative ratios, requiring anabolism to produce building blocks. Moreover, rapid proliferative growth caused by oncogenic activity might generate harmful oxidative species (29). These results may perhaps explain the altered levels of intermediates of 1- Carbon metabolism, S-Adenosyl-Methionine (SAM) cycle, and glutathione (GSH) (Figs. 2B, 2H, 2I, and 2J).

### A panel of six metabolites is associated with metastatic papillary thyroid cancer

In the literature percentage of metastatic (local and distant) DTC is relatively low (∼5%), but the prognosis is poorer than those with non-metastatic DTC (20, 23). However, most cases submitted to thyroid dissection in our cohort were metastatic (Fig. 3A). As there is a great need for reliable biomarkers for the metastatic potential of this tumor subset (30), we used such cohort composition in our favor to identify specific biomarkers that could help determine if a primary tumor was likely to metastasize to local lymph nodes or distant organs. We purposely removed the two samples from this analysis classified as FTC to avoid any noise in the classification. Therefore, we analyzed non-metastatic (n=6) and metastatic (n=11) PTC patient samples.

**Figure 3:**
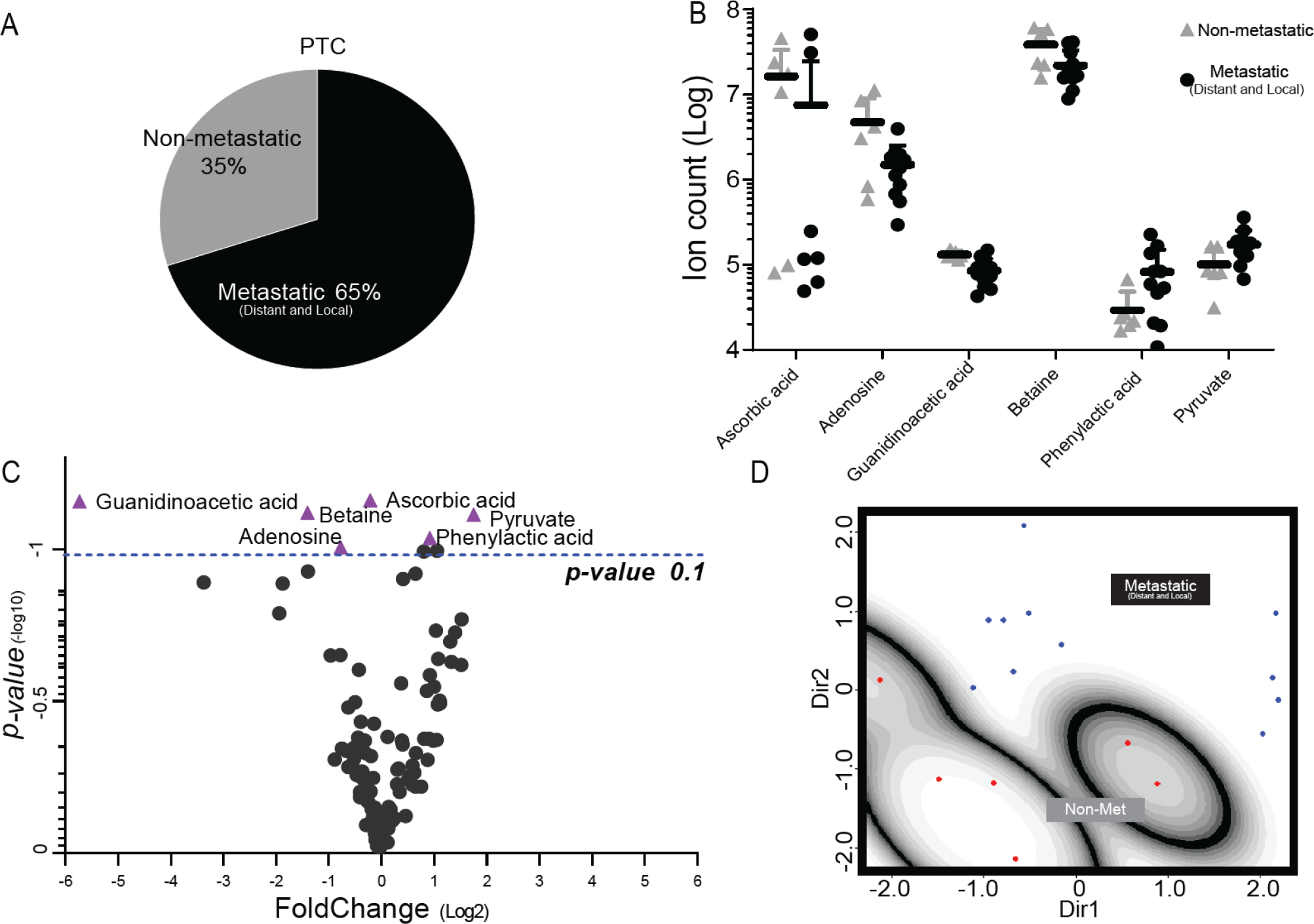
Six metabolites can help to identify metastatic papillary thyroid cancer. (A) Distribution of non-metastatic and metastatic (local and distant) PTC patients. (B) and (C) Most altered metabolites in metastatic PTC (*p*-value ≤0.1). (D) mclust projection showing the isolation of metastatic samples in a single group.

The ordinary logistic regression model was applied, evidencing six biomarkers with a *p*-value lower than 0.1: Adenosine, ascorbic acid, betaine, guanidinoacetic acid, phenylacetic acid, and pyruvate (Figs. 3B and 3C). None of the metabolites were found below the level of the Bonferroni-based cutoff (adjusted *p*-value≤0.0004545); however, we must consider this an extremely stringent criterion because of the much higher number of biomarkers compared to the number of samples. Nevertheless, the positive coefficients indicated by a non-adjusted *p*-value (*p*-value≤0.1) presented a positive correlation between these six metabolite levels and metastatic PTC tumors.

These six metabolites were considered for mclust analysis followed by 10-fold cross-validation (Fig. 3D). All metastatic PTC patients were classified as a single group (error rate 0.1176 with SE=0.0667). Notably, the 95% confidence interval excludes 0.5, indicating that this classification is statistically significant and that procedure abovementioned is robust enough to distinguish non-metastatic from metastatic PTC samples in this set.

### High nuclear and mitochondrial DNA mutation burden and shorter telomere length correlate to metastatic PTC

Ten PTC tumors with matched normal samples were randomly selected and submitted to whole-genome sequencing (WGS) with an average depth of 60x coverage. Four of these tumors were non-metastatic, and six were metastatic (local and distant metastasis) (Fig. 4A). The overall nuclear DNA (nDNA) mutation burden in PTC was low, generally less than 0.2 mutations per Mb (Fig. 4B). However, five PTC samples presented a nDNA mutation burden above this threshold. Four of them are metastatic, which might suggest a link between high nDNA mutation burden and metastatic potential in PTC (Supplementary Fig. S2A).

**Figure 4:**
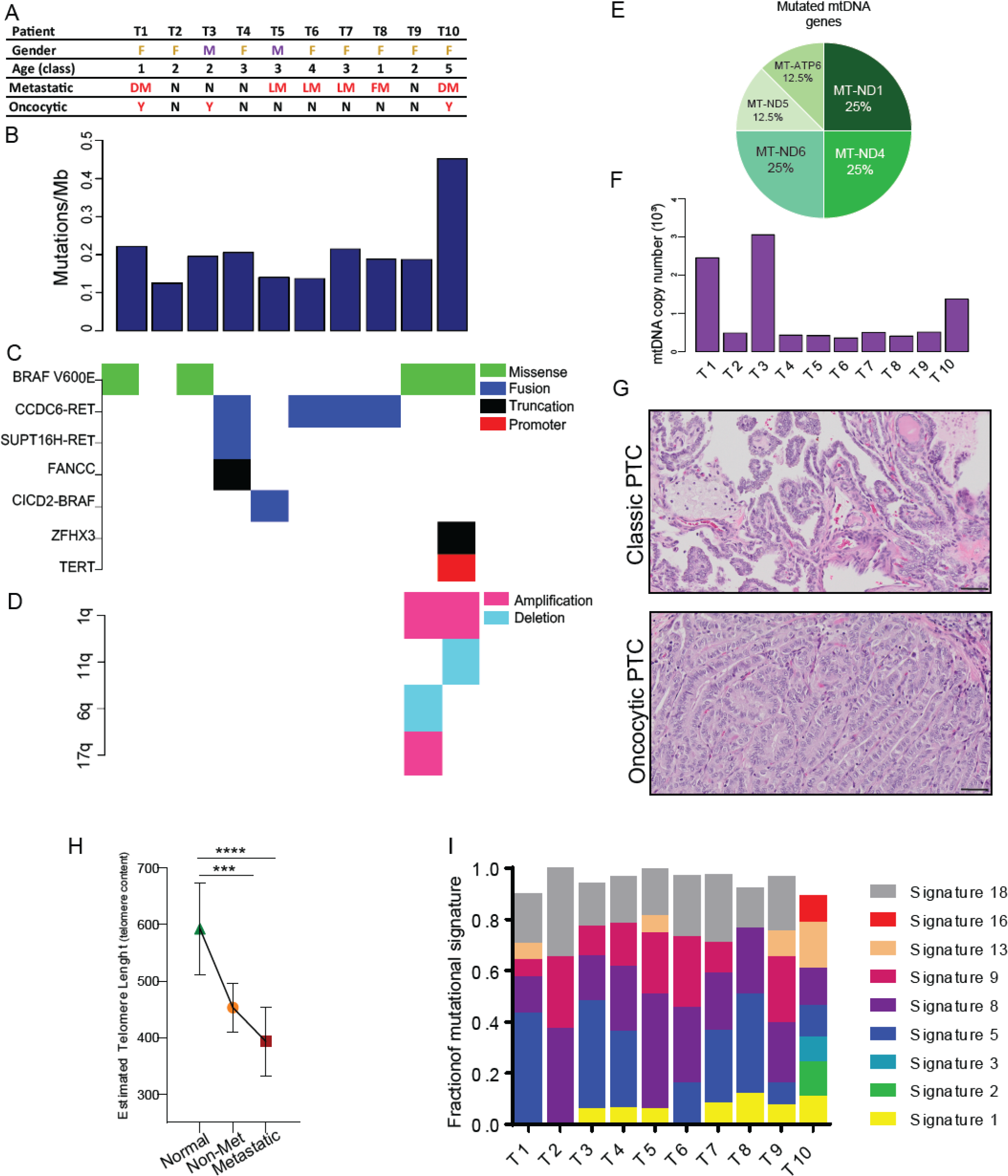
**Whole-genome sequencing of primary cancers suggests genetic features associated with metastatic PTC**. (A) General information and tumor characterization per patient across the cohort. Female and Male (F and M); Normal samples (N); Distant Metastasis (DM); Local Metastasis (LM). Age class (years): 1- (20-30); 2 - (31-40); 3 - (41-50); 4 - (51-60); 5 - (61-70). (B) Histogram presenting nuclear DNA mutation burden per sample. (C) Driver mutations in PTC. (D) Chromosome alterations. (E) Distribution of mtDNA mutations found in the genes that encode proteins of the mitochondrial electron transport chain. (F) Histogram presenting mtDNA copy number per sample. (G) H&E of scanned slides showing the histology of PTC classic type and PTC with oncocytic alterations (Scale bar 50µm). (H) Estimated telomere length in normal, non-metastatic, and metastatic PTC (I) COSMIC mutational signatures in PTC.

We identified seven driver mutations in this set of PTC samples (Fig. 4C). Five of these tumors have a BRAF mutation, four have the hotspot V600E mutation, and one has a CICD2-BRAF fusion. The structural variant analysis identified the other four thyroid tumors with inversions that resulted in CCDC6-RET fusions. While the breakpoints of the four inversions were all different, they all occur within intron-1 of CCDC6 and intron-11 of RET, resulting in the same fusion protein. Interestingly, our classification model based on the metabolic profile could not distinguish between samples with BRAF or RET alterations (Supplementary Figs. S2B and S2C). Although BRAF^V600E^ presented some association with more aggressive PTC elsewhere (31, 32), we could not find any correlation between this mutation and gain of aggressiveness in our cohort, as also reported by other groups (33).

Only two tumors showed meaningful copy number alterations. The tumor sample T9 had chromosome 1q duplication, 16q deletion, and 17q duplication (Fig. 4D). The other tumor sample (T10) had chromosome 1q duplication and 11q deletion.

The WGS average depth of coverage allows us to determine the mtDNA mutations by using GATK and Mutect2 (see Methods) and found missense and truncated mitochondrial gene mutations in six PTC samples (Supplementary Table S2). Most mtDNA mutations were found in genes that encode proteins from the electron transport chain (ETC) complex I, mainly in MT-ND1, MT-ND4, and MT-ND6; these alterations were also reported in previous articles (Fig. 4E) (34, 35). Metastatic PTC samples had a higher allelic frequency of mtDNA mutations (Supplementary Fig. S2D), suggesting that a high mtDNA mutation burden might be associated with metastatic risk.

The histopathology analysis of the samples with higher mtDNA copy numbers (namely, T1, T3, and T10) yielded PTC with oncocytic features (Figs. 4F and 4G), with an ubiquitous BRAF V600E mutation. Interestingly, one of the tumor samples (T10) had several mutations in mtDNA: MT-ND1, MT-ND4, and MT-ATP6. The mutation in MT-ND1 has an allele frequency of 0.83, demonstrating that this mutation is homoplastic, a feature found only in this tumor sample.

We also estimated the telomere length of all samples, including the normal thyroid tissue (Fig. 4H). The estimated telomere length of normal tissue compared with the tumor samples, metastatic or not, is much longer. Nonetheless, a clear trend indicates that metastatic PTC has a shorter telomere length than non-metastatic tumors.

Despite the absence of a specific gene mutation that could predict a more aggressive PTC, these results suggest that some genetic features, such as high nuclear and mitochondrial DNA mutation burden and shorter telomere length, are associated with metastatic PTC phenotype.

### Papillary thyroid cancer mutational signatures

The genetic sequencing of PTC samples presented nine different mutational signatures among the patients (Fig. 4I). The most common mutational signature is #8, which is present in all samples and is associated with possible exposure to radiation, one of the leading causes of thyroid cancer (36, 37). The second most common mutational signature is #18, found in nine patients. Curiously, signature #18 is linked to colibactin exposure. Colibactin is a mutagenic agent secreted by *E.coli*, which is highly associated with genotoxic colon cancer signatures but rarely reported in thyroid cancer (38). Additionally, the enrichment of signatures #8, #9, and #18 correlate with a substantial amount of C>A mutations in this cohort, a pattern of mutations associated with damage caused by reactive oxygen species (ROS) (39).

The mutational signatures #1 and #5 were found in most malignant tumor samples. These signatures correlate with the age at the time of cancer diagnosis and hence the nDNA mutational burden. However, in this cohort, the correlation between signatures #1 or #5 with nDNA mutation burden or age was not observed (Supplementary Figs. S2E and S2F).

The mutational signatures and the mutated gene pattern did not distinguish an aggressive PTC. Nonetheless, one patient sample (T10) presented the most advanced diseases with local and distant metastases. This patient also has a distinct characteristic presenting seven of nine (78%) listed signatures (Fig. 4I). This is the only sample demonstrating mutational signatures linked to age, smoking, and high numbers of insertions-deletions mutations (signatures #2, #3, and #16). All these features are present in this sample presumably because it belongs to the oldest patient, a former smoker, presenting an increased number of insertions-deletions (indels) and the highest nDNA mutation burden (Figs. 4B, 4D and Supplementary Table S1). Moreover, the tumor sample T10 is the only sample that presents three potential driver mutations: BRAF, ZFHX3, and TERT promoter (Fig. 4C). This sample had a high mtDNA burden, and it was the only sample with a homoplastic mutation (in MT-ND1 gene). Furthermore, the T10 patient also presented multifocal nodules and oncocytic sites in both thyroid glands. All these features suggest a history of high intratumor heterogeneity (ITH) that might perhaps play a role in high-risk PTC and deserve to be studied.

### Papillary thyroid cancer tumors present high metabolic, transcriptional, and immunogenic, but not genetic ITH

Intra-tumor genetic and non-genetic heterogeneity are established cancer hallmarks (40, 41), which often compromise therapy efficacy (42). We examined the extent of genetic and non-genetic ITH in two female patients (P1 and P2) of the same age and carrying PTC large enough for multi-omics profiling from multiple geographically distinct regions from their primary tumors. We selected four spatially well-separated areas from each primary tumor. We performed metabolomic, whole-exome sequencing, and RNA sequencing of these tumor regions and matched normal thyroid tissues (Fig. 5A).

**Figure 5:**
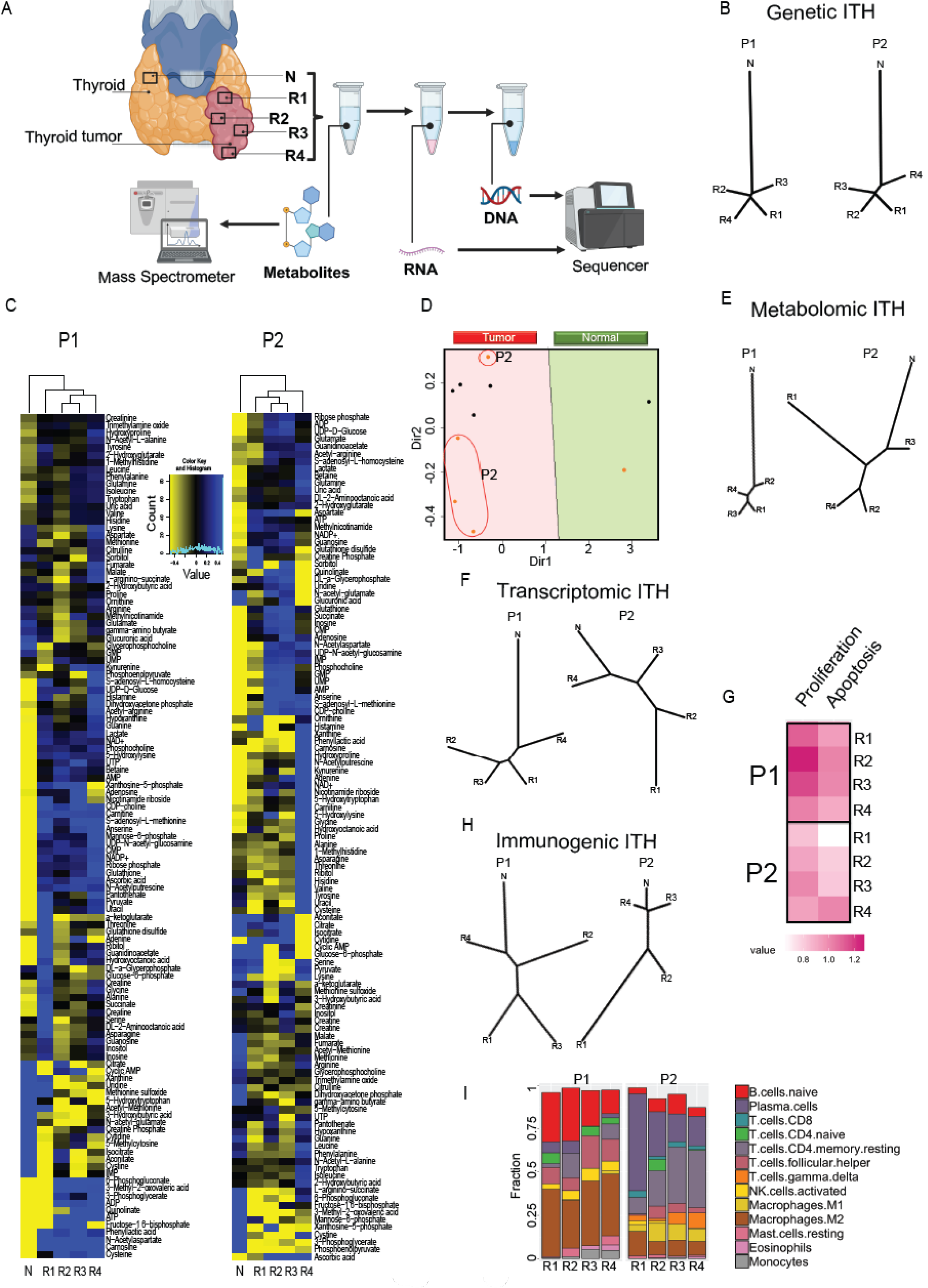
ITH in Papillary thyroid cancer. (A) Step-by-step genetic and non-genetic PTC intratumor heterogeneity determination. (B) Dendrogram of genetic ITH from patients 1 (P1) and 2 (P2). N represents normal thyroid tissue, and Rs (R1-R4) represent different pieces of PTC cancer tissues. (C) Metabolic heat maps, (D) principal component analysis, (E) and dendrogram show the distinct metabolic profile in different pieces of cancer in P2. (F) Dendrogram of transcriptional ITH. (G) Heat map of the proliferative score of cancer samples. (H) Dendrogram of Immunogenic ITH (I) Compositions of immune cells type infiltrated in cancer tissue.

Both patients had low somatic mutation burden and low genetic heterogeneity; most somatic mutations were ubiquitous and detected in all regions within the tumor (Fig. 5B), a pattern consistent with other reports (43).

In contrast, as reported in other tumors, ITH and divergence from the matched normal tissues were considerably higher at the transcriptomic, metabolic, and immunogenic levels (44). Tumor samples clustered separately from the normal tissues at the metabolite levels (Fig. 5 C), as observed previously (Fig. 1G). However, the metabolic ITH was considerably higher in P2, as shown by clustering and dendrograms (Figs. 5D and 5E). Moreover, the transcriptional ITH was observed to be increased in P2 (Fig. 5F). In addition, P2 presented a more conspicuous heterogeneous pattern in the proliferative score based on gene expression patterns (Fig. 5G). The heterogeneous metabolic and transcriptional profile of P2 reflects its tumor phenotype, a mixture of classic papillary type, follicular type, oncocytic features, and dense lymphocytic infiltration [7 of 12 metastatic lymph nodes (58%)]. On the other hand, P1 presents a non-metastatic classic type of papillary carcinoma. All the tumor samples from both patients presented very distinct immune cell profiles, which suggests a high immune ITH (Fig. 5H). Also, there was considerable variation between the patients in their proportional abundance of different immune cell types (Fig. 5I), suggesting that the immune microenvironment might differ among PTC samples.

Despite a small sample set, these results support a model that, while genetic variations are minor among the thyroid cancer subtypes, non-genetic heterogeneity at the metabolic, transcriptional, and immune levels within and between tumors, especially in metastatic PTC cases, was substantial.

## DISCUSSION

The potential for overdiagnosis and/or overtreatment in patients with thyroid cancer remains a critical topic for clinical research (8, 9, 45, 46). It is clear that DTC requires a more comprehensive procedure to identify patients that should or not be subjected to more invasive therapies. Another challenge in thyroid cancer translational research is the earlier identification of a potential metastatic PTC, which comprise ∼30% of all cases in the SEER database. Overall, the ten-year survival of thyroid cancer patients is ∼ 98%, but those few patients with local and distant metastases are associated with a very poor prognosis (11, 47). Unfortunately, efforts to find a mutational profile that predicts high-risk PTC, such as ThyroSeq (18) and Veracyte(17), are generally insufficient and may need to be combined with other diagnostic methods (16, 23). Considering that FNA aligned with cytopathologic analysis is inconclusive in many cases and the lack of data that can be used as a predictor of more aggressive thyroid cancer, we showed that an integrated analysis between non-genetics (metabolomic and cytopathology) and genetic biomarkers (RNA and DNA genome sequence) have the potential to refine the diagnosis and management of DTC.

The DTC metabolic profile is markedly different from normal thyroid tissue, as we showed, which is enough to identify thyroid cancer tissue, even when other conventional methods are inconclusive. Targeted and untargeted studies of thyroid cancer have suggested several potential metabolic signatures (48–50). Previous reports have proposed using metabolite levels as a potential diagnostic tool (51). Most of these metabolites, such as lactate, glucose, glutamine, asparagine, and choline, were indeed altered in our metabolic profile (52). However, the results presented here showed that other metabolites such as aconitate, glycine, inosine, isoleucine, proline, and taurine could be more efficient in distinguishing normal from cancer thyroid tissues, which might improve diagnostic accuracy.

Our study also showed that some metabolite levels are altered in metastatic PTC: adenine, ascorbic acid, betaine, guanidoacetic acid, phenylacetic acid, and phenylpyruvate. However, this phenotype must be associated with other cancer features to generate a more reliable distinction between low- and high-risk thyroid cancer. In this particular case, we suggest associating metabolomics with genomic data. Moreover, we might consider more than only BRAF mutations as predictors of poor prognostics (24, 43, 53). In the cancers with BRAF mutations in our cohort, 50% were metastatic, while the ones with RET fusions were largely metastatic (80%). Notably, we could not distinguish BRAF mutations and RET fusions at the metabolic level, presumably because they may redundantly trigger the same downstream signal transduction pathway(s). As such, both mutations were mutually-exclusive, as shown here and consistent with reports from other investigators (54–56). Taken together, this body of work suggests that both of these molecular alterations may ultimately lead tohigh-risk PTC generation. However, a major limitation of this discovery work is that our tentative hypothesis-generating conclusions are based on a relatively small patient cohort. Future studies of patients withPTC presenting with distant metastasis might shed further light on this open question.

Equally important, other alterations must be further investigated as potential high-risk PTC predictors, such as telomere length, nDNA and mtDNA mutation burden, and ITH. Our data showed a good correlation between shorter telomere length and cancer risk. Again, despite the relatively small patient cohort, we observed a high nDNA and mtDNA mutational burden and ITH in high-risk PTC when we contrasted multiple primary tumor regions of patients with non-metastatic versus metastatic PTC.

However, we should highlight that although the methods here were carefully applied, the conclusions of this study have inherited limitations due to its relatively small number of samples and its descriptive nature, which allows us only to suggest that these alterations could favor the identification of a metastatic PTC. Ideally, our results would need to be validated with a larger patient cohort.

This study showed that our classification model could distinguish cancer and normal thyroid tissue. This approach would be beneficial when the cytopathologic report presents indeterminate results and should help the physician to manage the patient, such as deciding whether to submit the patient to surgery, the extension of this surgery, and/or radioiodine therapy application. We also showed that multi-omics analysis considering telomere length, high nDNA and mtDNA mutation burden, and high ITH might help prematurely identify high-risk PTC. To conclude, we demonstrate that new research avenues in thyroid cancer may still have important implications for the diagnosis, treatment, and prognosis of DTC patients.

## METHODS

### Thyroid tissue extraction

The tissue was fractioned and classified by an expert dedicated pathologist. The DTC samples were picked from four equidistant sites to determine intratumor heterogeneity features. Paired normal and tumor thyroid tissue samples were fresh-frozen, unidentified, and maintained by the Cancer Institute of New Jersey Biospecimen Repository Service (CINJ-BRS) under the auspices of IRB-approved protocol. CINJ-BRS numbered and linked the stored tissue with its specific surgical report removing identification. The samples were obtained from CINJ-BRS after the protocol above and processed as indicated by the following protocols.

### Histology

A sample of each piece of tissue collected was fixed overnight in formalin 10% and then transferred to 70% ethanol for paraffin-embedded sections. The paraffin blocks were cut and mounted on slides. The slides were deparaffinized, rehydrated, and then submitted to hematoxylin-eosin staining.

### Tissue pulverization

A total of 25-30 mg of frozen thyroid tissue (duplicates of non-normal and tumor samples per patient) were weighed and added to a 2 mL round bottom microtube with a – 80° C cold Yttria Grinding Ball per tube. The tissues were pulverized by using a Retsch CryoMill following three alternating cycles at 5 Hz for 2 min and 25 Hz for 2 min.

### Extraction of polar metabolites from human thyroid tissue

To each 2 mL microtube with 25-30 mg of frozen pulverized tissue, a buffer volume equivalent to 20X the sample weight in μL was added. The extraction buffer was 40:40:20 (v/v/v) methanol:acetonitrile:water with 0.1 M of formic acid. After adding the buffer, the sample was vigorously vortexed and incubated on crushed ice for 10 minutes. The samples were then vortexed again and centrifuged for 10 min at 16,000 g at 4°C. The supernatant A was collected and saved, and the pellets were submitted to re-extraction following the same procedures which generated the supernatant B. Supernatant A and B were mixed and transferred to a clean 1.5 mL microtube with the appropriate volume of 15% NH4CO3. The samples were stored in a -80°C freezer until analysis by LC-MS.

### LC-MS analysis for polar metabolites

The LC-MS method involved hydrophilic interaction liquid chromatography (HILIC) coupled with electrospray ionization to the Q Exactive PLUS hybrid quadrupole-orbitrap mass spectrometer (Thermo Scientific). The LC separation was performed on an XBridge B.E.H. Amide column (150 mm × 2.1 mm, 2.5 μm particle size, Waters, Milford, MA) by using a gradient of solvent A (95%/5% H2O/ acetonitrile with 20 mM ammonium acetate and 20 mM ammonium hydroxide, pH 9.4), and solvent B (20%/80% H2O/ acetonitrile with 20 mM ammonium acetate and 20 mM ammonium hydroxide, pH 9.4). The gradient was 0 min, 100% B; 3 min, 100% B; 3.2 min, 90% B; 6.2 min, 90% B; 6.5 min, 80% B; 10.5 min, 80% B; 10.7 min, 70% B; 13.5 min, 70% B; 13.7 min, 45% B; 16 min, 45% B; 16.5 min, 100% B; 22 min, 100% B. The flow rate was 300 μL/min. The injection volume was 5 μL, and the column temperature was 25°C. The MS scans were done in both positive and negative ionization mode with a mass resolution of 70,000. The automatic gain control (AGC) target was 3e6. The maximum injection time was 50 ms. The scan range was 75-1,000. The metabolite features were extracted in MAVEN (57) with a mass accuracy window of 5 ppm. The compound identification is based on accurate mass and retention time matching our custmized in- house metabolite library (58).

### DNA and RNA extraction and processing

A total of 20-25 mg of frozen pulverized tissue was submitted to DNA extraction following DNeasy Blood and Tissue Kit (Qiagen ID:69504) protocol. For the ITH co-extraction of metabolites, DNA and RNA were extracted using the remaining pellet of tissues from the extraction of the metabolites. After initial quality checks of the raw RNA sequencing reads by using FastQC (v0.11.7) and removal of any low-quality reads, STAR aligner (v 2.6.0c) (59) was used to map the remaining reads onto the human genome (GRCh38). RSEM (v1.3.1) (60) was used for transcript quantification, and log2 (TPM+1) (Transcripts per million) values were reported for different tumor regions and also matched non-malignant regions. ESTIMATE (61) was used for predicting tumor purity and the presence of stromal/immune cells in tumor tissues. Multiregional tumor trees at transcriptomic levels were constructed for every patient with RNA expression data for all genes across different regions, an approach similar to that used at the genomic level. Manhattan distance was computed between all regions of a patient sample by using log2 (TPM+1), then unrooted dendrograms were drawn by using this distance metric. Proliferation (PI) and apoptotic (AI) indices were calculated a similar approach by considering 124 proliferation-associated genes (62) and six apoptosis-related genes. For each gene, Z-scores corresponding to its expression in tumor regions was calculated based on the mean and standard deviation of its expression in the non-malignant samples. Then proliferation index (PI) and apoptosis index (AI) were defined as mean z-scores of all proliferation and apoptosis genes, respectively.

### Whole genomic sequencing analysis

Mutect2 from the GATK (63) is carried out to analyze the point mutations and small indels by considering the tumor-normal pair mode with the GRCh38 genome. The corresponding results are annotated by SnpEff (63) and then filtered with a series of conditions: i). “FILTER” column as “PASS”; ii). tumor sample depth, REF + ALT > 10 & REF + ALT < 200; iii). normal sample depth, REF + ALT > 8 & REF + ALT < 200; iv). tumor AF > 0.2; v). normal AF < 0.05; and vi). SNVs only from 1– 22, X, Y, and MT chromosomes. The mutation signatures are analyzed by deconstructSigs (64) with the filtered SNVs as input. Manta analyzes the structural variants (SVs) (65). The filter conditions are applied to the original outputs: i). “FILTER” column as “PASS”; ii). SOMATICSCORE > 40. The copy number variations are investigated by using FACETS (66).

### Mitochondria DNA analysis

Ten tumor-normal pairs of whole-genome sequencing data were used for somatic mutation calling. First, the sequencing reads were clipped with Trimmomatic (Trimmomatic, RRID:SCR_011848) 0.39 (67); then, the reads were mapped to reference GRCh38 by using Burrows-Wheeler Alignment tools (bwa 0.7.17- r1188) (68). The bam files (the mapped file format) were further sorted, mark-duplicated, base quality score recalibrated (BQSR), and indexed using samtools-1.3.1 5 and gatk-4.1.9.0. Mitochondria-mode of Mutect2 (gatk) served to call somatic variants. The variants were filtered by gatk FilterMutercCalls. We developed a complex filter for screening low confident variants by using allele fractions of alternate alleles (AF) in the tumor (AF>0.03) and normal (AF=0), non-coding removal, approximate read depth (DP> 2000), and log ten likelihood ratio score of variant existing versus not existing (TLOD > 10). The variants were annotated by using Ensembl Variant Effect Predictor (VARIANT, RRID:SCR_005194; VEP 101.0) (69). The mtDNA copy number was estimated by the total number of reads mapping to the mitochondrial genome divided by the total number of reads and multiplied by the factor (6x10^9/16x10^3), which is the size of the diploid genome and the mitochondrial genome.

### Telomere length estimation method

We used TelomereHunter (70) to estimate telomere content from human WGS data considering default settings (44). Telomere content = Intra telomeric reads * 10^6^/ total reads with telomeric GC.

### Somatic mutation calling in ITH samples

FastQC (v0.11.7) was used for initial quality checks, and low-quality reads and PCR duplicates were removed. Next, we used BWA-mem (71) (v0.7.17-r1188) to map the reads onto human genome (GRCh38), and call variants by using varScan2 (72) (mapping quality > 40, base quality > 20). Only ‘high confidence’ somatic variants with tumor allele frequency > 5% at least in one tumor region and normal allele frequency < 1% were selected. For each somatic variant deemed as a high confidence variant in at least one tumor region, we queried the corresponding base position in other tumor regions in that tumor specimen. It was included if reads supported the variant allele with mapping quality > 20, base quality > 25, and variant allele frequency > 2%. All identified somatic mutations were annotated with SnpEff (63)(v4.3t). Missense, nonsense, frameshift, or splicing mutations in known COSMIC cancer genes with high-predicted impact were marked. The dendrograms were generated with the approach described (44) in the previous paper from variant allele frequency of variations identified in different regions.

### Mutational signatures

Contexts of somatic point mutations were used to draw inferences about their likely etiologies (73). We used deconstructSigs (39, 64) to identify patterns of mutational signatures on somatic variants.

### Immune cell infiltration

Immune cell infiltrations were inferred from molecular signatures of immune cell types. ESTIMATE (v1.0.13) (61) was used to predict the immune infiltration level in tumor tissues. CIBERSORT (74) was used to estimate the abundance of different immune cell populations from expression data. Standard LM22 signature gene file and 1000 permutations were used to calculate deconvolution p values. Similar to genomic and transcriptomic data, multiregional tumor trees were made for every patient to infer immunogenic heterogeneity (iITH) with immune cell proportions from CIBERSORT. Manhattan distance was computed between all regions of a patient sample using immune cell proportions datasets independently, and separate unrooted dendrograms were drawn by using respective distance metrics.

### Pathways impact analysis and metabolites classification

To evaluate the impact of the metabolic alterations found in tumor versus normal thyroid tissues, we submitted the raw data obtained from the metabolic extraction on MAVEN (ion counts) to log transformation by using the web-based software MetaboAnalyst 5.0 (75). The transformed data were used to obtain the class of the metabolites significantly altered and the impact of these alterations on tumor thyroid tissues (76).

### Graphs and figures

All charts and statistics presented in this article were built using GraphPad Prism 9 (GraphPad Prism, RRID:SCR_002798) and RStudio 1.1453. The cartoon figure 5A was created with BioRender (Biorender, RRID:SCR_018361). All graphs, pictures, and cartoons were assembled by using Adobe Illustrator 26 (Adobe Illustrator, RRID:SCR_010279).

### Predictive model and statistical analysis

First, the biomarker levels of the 110 metabolites were transformed by using a log2 transformation. Since the number of markers is zero, a small amount, 100, was added to all the biomarkers before taking logs. Thus, the transformation is given by y=log2(x+100). As a first step in constructing a predictive model, we used ordinary logistic regression to assess the relationship of each predictor to the sample type. Then the biomarkers that presented p-values smaller than 0.20/110 = 0.0018 (Bonferroni adjustment) were selected as predictors unless mentioned differently. The numbers and kinds of predictors are different depending on the comparison. The Lasso regression (25) was applied to find a subset of biomarkers that effectively predict the status of the sample. Additionally, we used the “profL1” function of the “penalized” R package to select, via cross-validation, the optimal tuning parameter. Then we used the “penalized” function to carry out the lasso regression.

### Study approval

All patients consented to submit their samples to this protocol in writing. All tissue samples were acquired by partial or total thyroid dissection per the approved protocol (CINJ 001724 – Pro20170001082) and Institutional Review Board (IRB) approval at Rutgers Cancer Institute of New Jersey, New Brunswick, NJ, USA.

## Author Contribution

ECL performed experimental work, data analysis, and data interpretation and wrote the paper. E.W. is the leading investigator that conceived the project, supervised research, and edited the paper. A.Sa assisted with the clinical protocol, sample preparation, and data analysis. DM developed the algorithmic classification model and assisted with the statistical analysis. HK, FS, SL, YC, and CSC processed and analyzed the whole genome sequencing data. A.Sh and SD processed the ITH samples and ITH DNA and RNA sequence data. JN assisted with sample preparation. XS performed metabolomic processing and analysis. GR and SG classified the tumor samples and evaluated the surgical reports of the patients. DM, JYG, MG, MI, GR, EL, TLS, ST, SG, XS, SD, RP, WA, and CSC interpreted the data and edited the paper.

## Data Availability

All data produced in the present work are contained in the manuscript

## Acknowledgments

This work was supported by R01 CA243547 to EW, ECL, and SG; Robert Wood Johnson Foundation (Project number #73711). The Biospecimen Repository and Histopathology Service Shared Resource from the Cancer Institute of New Jersey provided all the specimens and associated services (P30CA072720-5919); Biometrics Shared Resource (P30CA072720-5918); Metabolomics Shared Resource (P30CA072720-5923), Bioinformatics Shared Resources (P30CA072720-5917).

## SUPPLEMENTARY FIGURES AND TABLES

**Supplementary Figure S1:**
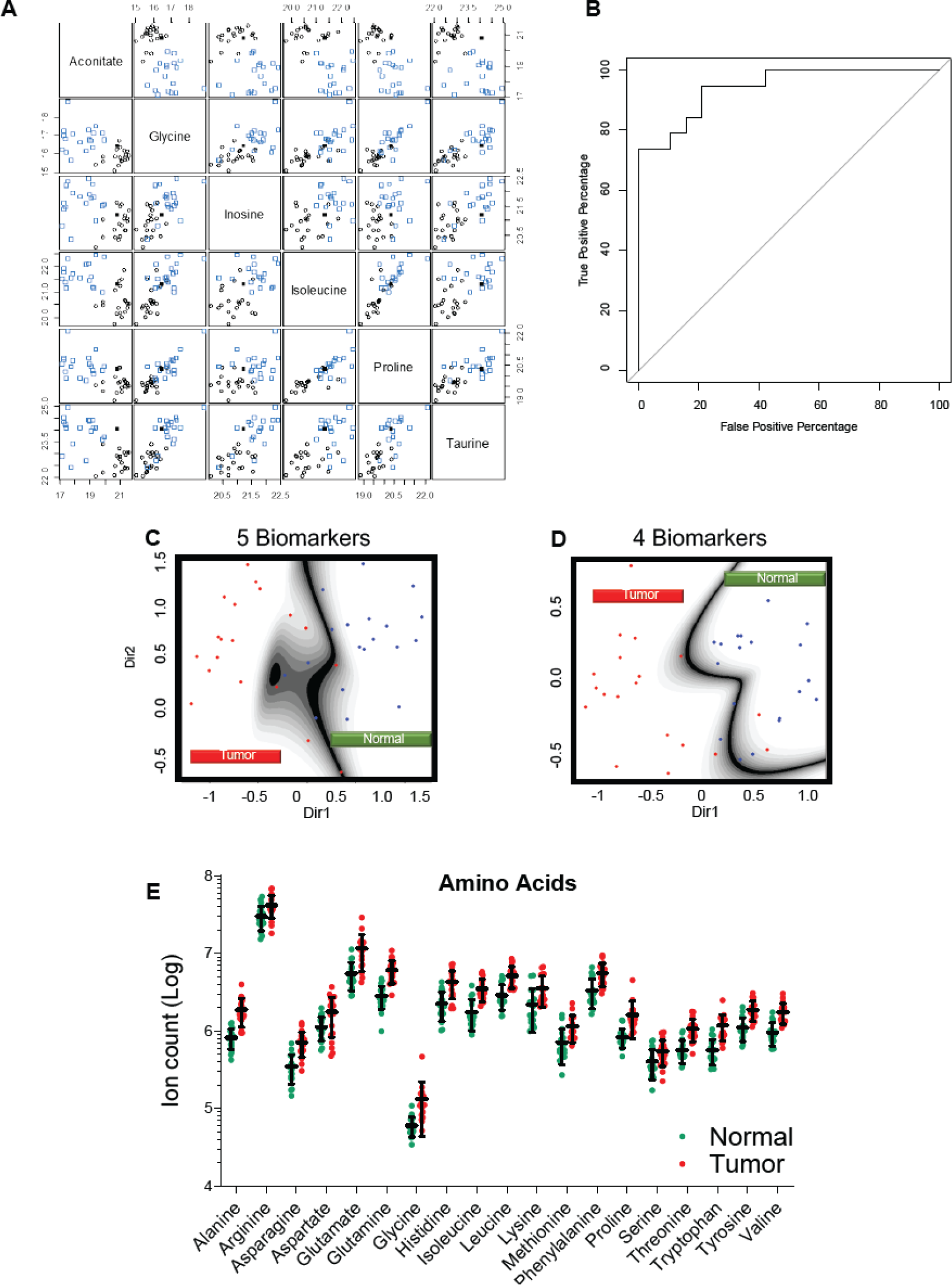
(A) Matrix of scatterplots of each of the seven biomarkers vs. the others, with “blue squares” indicating cancer samples and “black circles” normal samples. (B) The receiver operating characteristic curve (ROC) shows the predictive model’s power. mClust analysis considering (C) five (glutamate, leucine, proline, taurine, and threonine) and (D) four (glutamate, proline, taurine, and threonine) most altered metabolites. The cross-validated and standard error rates are 0.1579 - 0.055 for five biomarkers and 0.236 - 0.059 for four biomarkers. (E) Levels of proteogenic amino acids found in DTC patients’ samples.

**Supplementary Figure S2:**
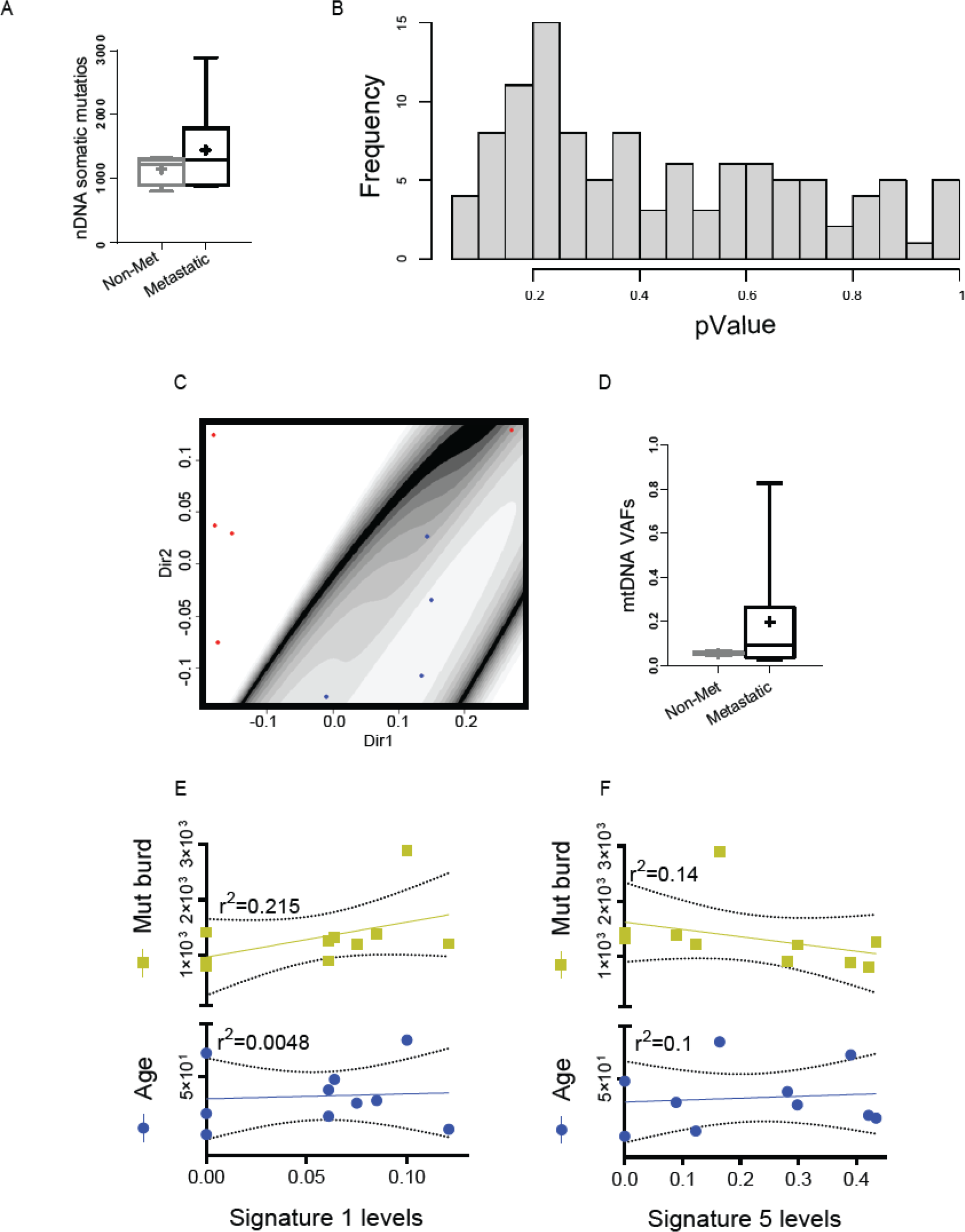
(A) Nuclear DNA mutation burden in metastatic and non-metastatic PTC. (B) Logistic regression analysis of BRAF mutation versus CCDC6-RET fusion comparison presenting the p- values per sample. The smallest p-value in this comparison was 0.0760 (thiamine), emphasizing no correlation between the metabolic profile and genetic alterations. (C) mClust analysis considering the six main altered metabolites comparing the groups in B. (D) mtDNA mutation burden in metastatic and non- metastatic PTC. (E) and (F) Correlation of nDNA mutation burden and age with mutational signatures 1 and 5, respectively.

**Supplementary Table 1:**
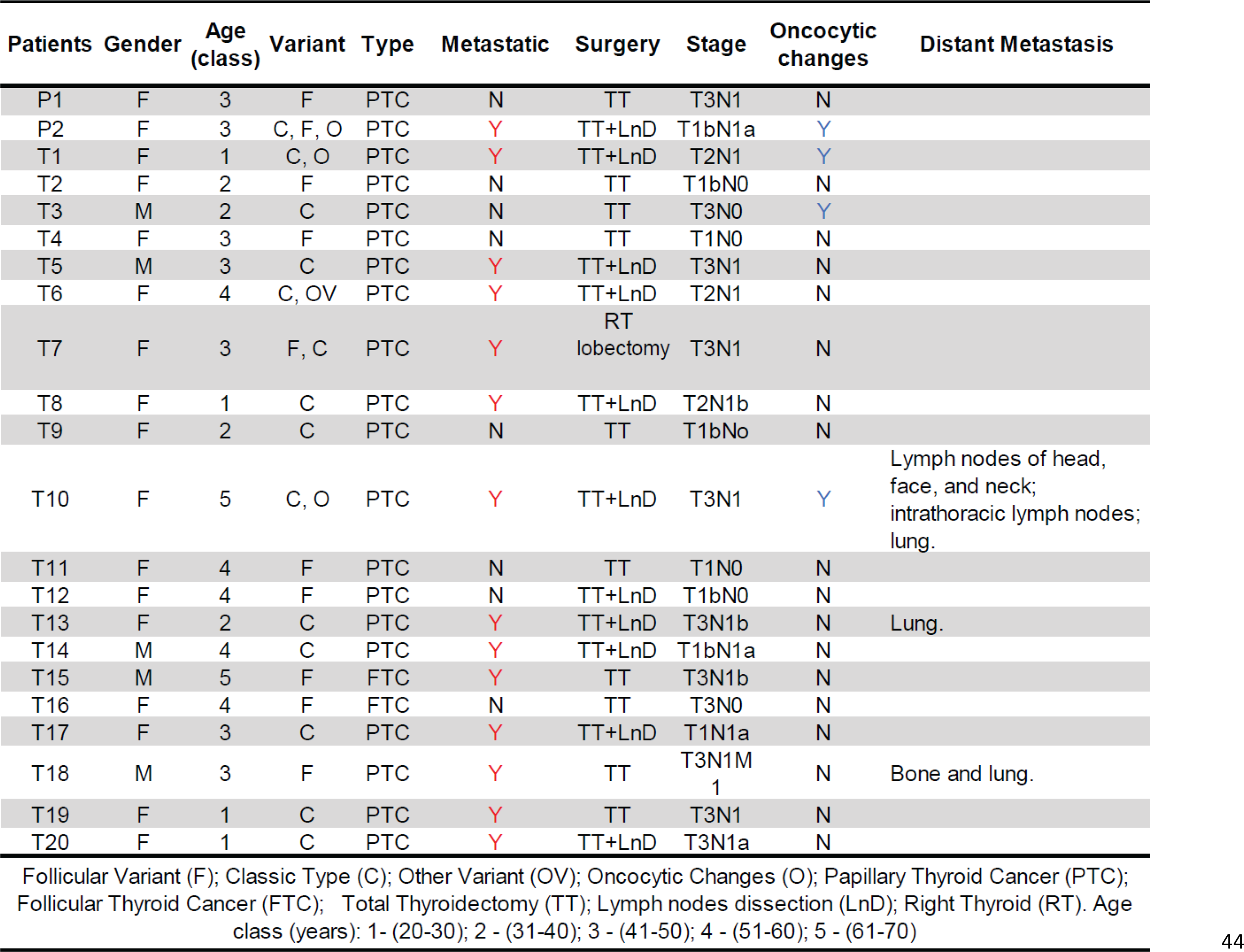
Patients information

| Patients | Gender | Age<br>(class) | Variant | Type | Metastatic | Surgery | Stage | Oncocytic<br>changes | Distant Metastasis |
| --- | --- | --- | --- | --- | --- | --- | --- | --- | --- |
| P1 | F | 3 | F | PTC | N | TT | T3N1 | N |  |
| P2 | F | 3 | C, F, O | PTC | Y | TT+LnD | T1bN1a | Y |  |
| T1 | F | 1 | C, O | PTC | Y | TT+LnD | T2N1 | Y |  |
| T2 | F | 2 | F | PTC | N | TT | T1bN0 | N |  |
| T3 | M | 2 | C | PTC | N | TT | T3N0 | Y |  |
| T4 | F | 3 | F | PTC | N | TT | T1N0 | N |  |
| T5 | M | 3 | C | PTC | Y | TT+LnD | T3N1 | N |  |
| T6 | F | 4 | C, OV | PTC | Y | TT+LnD | T2N1 | N |  |
| T7 | F | 3 | F, C | PTC | Y | RT<br>lobectomy | T3N1 | N |  |
| T8 | F | 1 | C | PTC | Y | TT+LnD | T2N1b | N |  |
| T9 | F | 2 | C | PTC | N | TT | T1bNo | N |  |
| T10 | F | 5 | C, O | PTC | Y | TT+LnD | T3N1 | Y | Lymph nodes of head,<br>face, and neck;<br>intrathoracic lymph nodes;<br>lung. |
| T11 | F | 4 | F | PTC | N | TT | T1N0 | N |  |
| T12 | F | 4 | F | PTC | N | TT+LnD | T1bN0 | N |  |
| T13 | F | 2 | C | PTC | Y | TT+LnD | T3N1b | N | Lung. |
| T14 | M | 4 | C | PTC | Y | TT+LnD | T1bN1a | N |  |
| T15 | M | 5 | F | FTC | Y | TT | T3N1b | N |  |
| T16 | F | 4 | F | FTC | N | TT | T3N0 | N |  |
| T17 | F | 3 | C | PTC | Y | TT+LnD | T1N1a | N |  |
| T18 | M | 3 | F | PTC | Y | TT | T3N1M<br>1 | N | Bone and lung. |
| T19 | F | 1 | C | PTC | Y | TT | T3N1 | N |  |
| T20 | F | 1 | C | PTC | Y | TT+LnD | T3N1a | N |  |

**Supplementary Table 2:** **mtDNA mutations**

| **Tumor ID** | **Position** | **Reference** | **Alteration** | **Mutation Type** | **Gene** | **Tumor AF (%)** |
| --- | --- | --- | --- | --- | --- | --- |
| **T1** | 13886 | T | C | Missense | MT-ND5 | 0.093 |
| **T2** | 10946 | A | AC | Frameshift | MT-ND4 | 0.068 |
| **T3** | 3982 | G | AC | Missense | MT-ND1 | 0.045 |
| **T6** | 14162 | G | AC | Missense | MT-ND6 | 0.177 |
| **T7** | 14527 | A | AC,ACC,ACCC | Frameshift | MT-ND6 | 0.363, 0.095, 0.024 |
| **T10** | 3565 | A | AC, ACC | Frameshift | MT-ND1 | 0.827, 0.029 |
| **T10** | 8872 | G | A | Missense | MT-ATP6 | 0.148 |
| **T10** | 10946 | A | AC | Frameshift | MT-ND4 | 0.037 |

Supplementary Table 2: mtDNA mutations
| Tumor ID | Position | Reference | Alteration | Mutation Type | Gene | Tumor AF (%) |
| --- | --- | --- | --- | --- | --- | --- |
| T1 | 13886 | T | C | Missense | MT-ND5 | 0.093 |
| T2 | 10946 | A | AC | Frameshift | MT-ND4 | 0.068 |
| T3 | 3982 | G | AC | Missense | MT-ND1 | 0.045 |
| T6 | 14162 | G | AC | Missense | MT-ND6 | 0.177 |
| T7 | 14527 | A | AC,ACC,ACCC | Frameshift | MT-ND6 | 0.363, 0.095, 0.024 |
| T10 | 3565 | A | AC, ACC | Frameshift | MT-ND1 | 0.827, 0.029 |
| T10 | 8872 | G | A | Missense | MT-ATP6 | 0.148 |
| T10 | 10946 | A | AC | Frameshift | MT-ND4 | 0.037 |

## REFERENCES

1. Xing M. Molecular pathogenesis and mechanisms of thyroid cancer. Nat. Rev. Cancer 2013;13(3):184–199.

2. Levi F. Cancer incidence in five continents, vol. VI. 1993:

3. Morris LGT, et al. The increasing incidence of thyroid cancer: The influence of access to care. Thyroid 2013;23(7):885–891.

4. Siegel RL, et al. Cancer Statistics, 2021. CA. Cancer J. Clin. 2021;71(1):7–33.

5. Chen AY, Jemal A, Ward EM. Increasing incidence of differentiated thyroid cancer in the United States, 1988-2005. Cancer 2009;115(16):3801–3807.

6. Kent WDT, et al. Increased incidence of differentiated thyroid carcinoma and detection of subclinical disease. CMAJ 2007;177(11):1357–1361.

7. Enewold L, et al. Rising thyroid cancer incidence in the United States by demographic and tumor characteristics, 1980-2005. Cancer Epidemiol. Biomarkers Prev. 2009;18(3):784–791.

8. Haugen BR, et al. 2015 American Thyroid Association Management Guidelines for Adult Patients with Thyroid Nodules and Differentiated Thyroid Cancer: The American Thyroid Association Guidelines Task Force on Thyroid Nodules and Differentiated Thyroid Cancer. Thyroid 2016;26(1):1–133.

9. Nabhan F, Ringel MD. Thyroid nodules and cancer management guidelines: comparisons and controversies. Endocr. Relat. Cancer 2017;24(2):R13–R26.

10. Nayar R, Ivanovic M. The indeterminate thyroid fine-needle aspiration: Experience from an academic center using terminology similar to that proposed in the 2007 national cancer institute thyroid fine needle aspiration state of the science conference. Cancer Cytopathol. 2009;117(3):195–202.

11. Ho AS, et al. Increasing diagnosis of subclinical thyroid cancers leads to spurious improvements in survival rates. Cancer 2015;121(11):1793–1799.

12. Davies L, Welch HG. Current Thyroid Cancer Trends in the United States. JAMA Otolaryngol. Neck Surg. 2014;140(4):317.

13. Bible KC, et al. 2021 American Thyroid Association Guidelines for Management of Patients with Anaplastic Thyroid Cancer. Thyroid 2021;31(3):337–386.

14. Cibas ES, Ali SZ. The 2017 Bethesda System for Reporting Thyroid Cytopathology. Thyroid 2017;27(11):1341– 1346.

15. Wang CCC, et al. A large multicenter correlation study of thyroid nodule cytopathology and histopathology. Thyroid 2011;21(3):243–251.

16. Iñiguez-Ariza NM, Brito JP. Management of Low-Risk Papillary Thyroid Cancer. Endocrinol. Metab. 2018;33(2):185–194.

17. Alexander EK, et al. Preoperative Diagnosis of Benign Thyroid Nodules with Indeterminate Cytology. N. Engl. J. Med. 2012;367(8):705–715.

18. Nikiforov YE, et al. Performance of a Multigene Genomic Classifier in Thyroid Nodules With Indeterminate Cytology: A Prospective Blinded Multicenter Study. JAMA Oncol. 2019;5(2):204–212.

19. Livhits MJ, et al. Effectiveness of Molecular Testing Techniques for Diagnosis of Indeterminate Thyroid Nodules: A Randomized Clinical Trial. JAMA Oncol. 2021;7(1):70–77.

20. Nixon IJ, et al. Management of Invasive Differentiated Thyroid Cancer. Thyroid 2016;26(9):1156–1166.

21. Weber T, Klar E. Minimal residual disease in thyroid carcinoma. Semin. Surg. Oncol. 2001;20(4):272–277.

22. Brito JP, et al. A Clinical Framework to Facilitate Risk Stratification When Considering an Active Surveillance Alternative to Immediate Biopsy and Surgery in Papillary Microcarcinoma. Thyroid 2016;26(1):144–149.

23. Janjua N, Wreesmann VB. Aggressive differentiated thyroid cancer. Eur. J. Surg. Oncol. 2018;44(3):367–377.

24. D’Cruz AK, et al. Molecular markers in well-differentiated thyroid cancer. Eur. Arch. Oto-Rhino-Laryngology 2018;275(6):1375–1384.

25. Hastie, T., Tibshirani, R. & Friedman J. The elements of statiscal learning. Springer Ser. Stat. 2009;26(4):505–516.

26. Scrucca L, et al. mclust 5: Clustering, Classification and Density Estimation Using. R J. 2016;8(1):289–317.

27. Fraley C, Raftery AE. MCLUST: Software for Model-Based Cluster Analysis. J. Classif. 1999;16(2):297–306.

28. Stephens FB, Constantin-teodosiu D, Greenhaff PL. New insights concerning the role of carnitine in the regulation of fuel metabolism in skeletal muscle. J. Physiol. 2007;581(2):431–444.

29. Cararo-Lopes E, et al. Autophagy buffers Ras-induced genotoxic stress enabling malignant transformation in keratinocytes primed by human papillomavirus (Cell Death & Disease, (2021), 12, 2, (194), 10.1038/s41419-021- 03476-3). Cell Death Dis. 2021;12(4). doi:10.1038/s41419-021-03564-4

30. Cabanillas ME, McFadden DG, Durante C. Thyroid cancer. Lancet 2016;388(10061):2783–2795.

31. Cohen Y, et al. BRAF mutation in papillary thyroid carcinoma. Hournal Natl. Cancer Inst. 2003;95(8):625–627.

32. Santoro M, Carlomagno F. Central Role of RET in Thyroid Cancer. Cold Spring Harb. Perspect. Biol. 2013;5(12):a009233.

33. Agrawal N, et al. Integrated Genomic Characterization of Papillary Thyroid Carcinoma. Cell 2014;159(3):676–690.

34. Chatterjee A, Mambo E, Sidransky D. Mitochondrial DNA mutations in human cancer. Oncogene 2006;25(34):4663–4674.

35. Yuan Y, et al. Comprehensive Molecular Characterization of Mitochondrial Genomes in Human Cancers[published online ahead of print: 2017]; doi:10.1101/161356

36. Alexandrov LB, et al. The repertoire of mutational signatures in human cancer. Nature 2020;578(7793):94–101.

37. Ron E, et al. Thyroid cancer after exposure to external radiation: A pooled analysis of seven studies. Radiat. Res. 1995;141(3):259–277.

38. Pleguezuelos-Manzano C, et al. Mutational signature in colorectal cancer caused by genotoxic pks + E. coli. Nature 2020;580(7802):269–273.

39. Alexandrov LB, et al. Signatures of mutational processes in human cancer. Nature 2013;500(7463):415–421.

40. Biswas A, De S. Drivers of dynamic intratumor heterogeneity and phenotypic plasticity. Am. J. Physiol. - Cell Physiol. 2021;320(5):C750–C760.

41. Hanahan D. Hallmarks of Cancer: New Dimensions. Cancer Discov. 2022;12(1):31–46.

42. McGranahan N, Swanton C. Biological and therapeutic impact of intratumor heterogeneity in cancer evolution. Cancer Cell 2015;27(1):15–26.

43. Agrawal N, et al. Integrated Genomic Characterization of Papillary Thyroid Carcinoma. Cell 2014;159(3):676–690.

44. Sharma A, et al. Non-Genetic Intra-Tumor Heterogeneity Is a Major Predictor of Phenotypic Heterogeneity and Ongoing Evolutionary Dynamics in Lung Tumors. Cell Rep. 2019;29(8):2164–2174.e5.

45. Brito JP, et al. Overdiagnosis of thyroid cancer and Graves’ disease. Thyroid 2014;24(2):402–403.

46. Lim H, et al. Trends in thyroid cancer incidence and mortality in the United States, 1974-2013. JAMA - J. Am. Med. Assoc. 2017;317(13):1338–1348.

47. Yang L, Shen W, Sakamoto N. Population-based study evaluating and predicting the probability of death resulting from thyroid cancer and other causes among patients with thyroid cancer. J. Clin. Oncol. 2013;31(4):468–474.

48. Khatami F, et al. Oncometabolites as biomarkers in thyroid cancer: a systematic review. Cancer Manag. Res. 2019;Volume 11:1829–1841.

49. Yao Z, et al. Serum metabolic profiling and features of papillary thyroid carcinoma and nodular goiter. Mol. Biosyst. 2011;7(9):2608–2614.

50. Xu Y, et al. Distinct Metabolomic Profiles of Papillary Thyroid Carcinoma and Benign Thyroid Adenoma. J. Proteome Res. 2015;14(8):3315–3321.

51. Khatami F, et al. Oncometabolites as biomarkers in thyroid cancer: A systematic review. Cancer Manag. Res. 2019;11:1829–1841.

52. Wishart DS, et al. Cancer metabolomics and the human metabolome database. Metabolites 2016;6(1). doi:10.3390/metabo6010010

53. Nikiforov YE, et al. Impact of mutational testing on the diagnosis and management of patients with cytologically indeterminate thyroid nodules: A prospective analysis of 1056 FNA samples. J. Clin. Endocrinol. Metab. 2011;96(11):3390–3397.

54. Stransky N, et al. The landscape of kinase fusions in cancer. Nat. Commun. 2014;5. doi:10.1038/ncomms5846

55. Yakushina VD, Lerner L V., Lavrov A V. Gene fusions in thyroid cancer. Thyroid 2018;28(2):158–167.

56. Cantwell-Dorris ER, O’Leary JJ, Sheils OM. BRAFV600E: Implications for carcinogenesis and molecular therapy.Mol. Cancer Ther. 2011;10(3):385–394.

57. Melamud E, Vastag L, Rabinowitz JD. Metabolomic analysis and visualization engine for LC - MS data. Anal. Chem. 2010;82(23):9818–9826.

58. Lu W, et al. Metabolite Measurement: Pitfalls to Avoid and Practices to Follow. Annu. Rev. Biochem. 2017;86:277.

59. Dobin A, et al. STAR: ultrafast universal RNA-seq aligner. Bioinformatics 2013;29(1):15–21.

60. Li B, Dewey CN. RSEM: Accurate transcript quantification from RNA-Seq data with or without a reference genome. BMC Bioinformatics 2011;12(1):1–16.

61. Yoshihara K, et al. Inferring tumour purity and stromal and immune cell admixture from expression data. Nat. Commun. 2013 41 2013;4(1):1–11.

62. Chen Z, et al. A murine lung cancer co-clinical trial identifies genetic modifiers of therapeutic response. Nat. 2012 4837391 2012;483(7391):613–617.

63. Cingolani P, et al. A program for annotating and predicting the effects of single nucleotide polymorphisms, SnpEff: SNPs in the genome of Drosophila melanogaster strain w1118; iso-2; iso-3. Fly (Austin). 2012;6(2):80.

64. Rosenthal R, et al. deconstructSigs: Delineating mutational processes in single tumors distinguishes DNA repair deficiencies and patterns of carcinoma evolution. Genome Biol. 2016;17(1):1–11.

65. Chen X, et al. Manta: Rapid detection of structural variants and indels for germline and cancer sequencing applications. Bioinformatics 2016;32(8):1220–1222.

66. Shen R, Seshan VE. FACETS: Allele-specific copy number and clonal heterogeneity analysis tool for high-throughput DNA sequencing. Nucleic Acids Res. 2016;44(16):1–9.

67. Bolger AM, Lohse M, Usadel B. Trimmomatic: a flexible trimmer for Illumina sequence data. Bioinformatics 2014;30(15):2114–2120.

68. Li H. Aligning sequence reads, clone sequences and assembly contigs with BWA-MEM[published online ahead of print: March 16, 2013];https://arxiv.org/abs/1303.3997v2. cited January 11, 2022

69. Howe KL, et al. Ensembl 2021. Nucleic Acids Res. 2021;49(D1):D884–D891.

70. Feuerbach L, et al. TelomereHunter - In silico estimation of telomere content and composition from cancer genomes. BMC Bioinformatics 2019;20(1):1–11.

71. Li H, Durbin R. Fast and accurate short read alignment with Burrows-Wheeler transform. Bioinformatics 2009;25(14):1754–1760.

72. Koboldt DC, et al. VarScan 2: Somatic mutation and copy number alteration discovery in cancer by exome sequencing. Genome Res. 2012;22(3):568–576.

73. Hu X, Xu Z, De S. Characteristics of mutational signatures of unknown etiology. NAR cancer 2020;2(3). doi:10.1093/NARCAN/ZCAA026

74. Gentles AJ, et al. The prognostic landscape of genes and infiltrating immune cells across human cancers. Nat. Med. 2015 218 2015;21(8):938–945.

75. Chong J, Wishart DS, Xia J. Using MetaboAnalyst 4.0 for Comprehensive and Integrative Metabolomics Data Analysis. Curr. Protoc. Bioinforma. 2019;68(1):e86.

76. Xia J, Wishart DS. Web-based inference of biological patterns, functions and pathways from metabolomic data using MetaboAnalyst. Nat. Protoc. 2011 66 2011;6(6):743–760.

